# From Seroprevalence to Measles Outbreak Risk: A Multicountry Epidemiological Proof of Concept

**DOI:** 10.64898/2026.07.28.26359095

**Authors:** Eran Kopel, Ravit Bassal

**Affiliations:** Tel Aviv University

**Keywords:** measles, seroprevalence, susceptibility, outbreak forecasting, vaccination, importation

## Abstract

**Background:** High national vaccination coverage may conceal age-specific and spatially concentrated measles susceptibility.

**Objective:** To assess whether published age-specific seropositivity results can be converted into a time-updated susceptibility profile that corresponds with subsequent measles incidence, while distinguishing susceptibility from infectious introductions and transmission conditions.

**Methods:** For Israel, published 2015 age-specific seropositivity estimates were mapped to monthly birth cohorts and projected to 1 March 2018, accounting for births, aging, maternal antibody, routine vaccination, vaccine effectiveness, and uncertainty in assay interpretation. The primary outcome was reported age-specific incidence during the 2018-2019 outbreak; national and Jerusalem District case burdens were secondary outcomes. Published evidence from the Netherlands, Czechia, and Australia was compared using a common framework covering age distribution, assay classification, vaccination, importation, spatial concentration, and transmission context.

**Results:** The estimated number susceptible in Israel on 1 March 2018 ranged from approximately 0.55 million (6.3% of the modelled population) to 1.92 million (22.4%), with a central estimate of 1.22 million (14.2%). Children aged <1 year had the highest central susceptible proportion (75.7%) and the highest later incidence (196.0 per 100,000). Jerusalem District accounted for 2,202 of 4,311 reported national cases, consistent with susceptibility concentrated in communities with lower first-dose coverage. The external comparisons showed that clustering amplified Dutch outbreak risk, survey design affected Czech estimates, and importation dominated Australian activity.

**Conclusions:** Published seropositivity can identify immunity gaps, but useful outbreak-risk assessment must also represent susceptible density and distribution, introduction pressure, and local transmission conditions. Although the model was not designed to compare alternative vaccination schedules directly, its identification of substantial susceptibility during early childhood provides epidemiological support for Israel’s recent decision to advance the second routine MMRV dose from 6 years to 18 months of age, thereby shortening the period during which young children remain dependent on single-dose protection.

## Introduction

Measles has a high transmission potential, and national averages of vaccination coverage or seropositivity may obscure epidemiologically important immunity gaps. Populations with apparently adequate national protection may still contain age groups, localities, or social networks in which susceptibility is concentrated. Following an infectious introduction, transmission is more likely to persist when susceptible individuals are numerous, geographically dense, and connected through repeated contact [1,2]. The relevant population measure is therefore not only the overall susceptible proportion, but also the age distribution, spatial density, and contact-network concentration of susceptible individuals.

Existing serological measurements are often separated in time from the outbreak period they are intended to inform. Their interpretation therefore requires cohort aging and explicit allowance for births, routine vaccination, and changing infant protection. We aimed to determine whether published age-specific seropositivity results could be converted into a time-updated susceptibility profile that corresponded with later age-specific measles incidence, and to identify which additional contextual factors were required to interpret outbreak occurrence across settings.

## Methods

### Study design and outcomes

We conducted a retrospective proof-of-concept analysis using published aggregate results and public demographic information. No individual-level serosurvey or surveillance records were accessed. For Israel, the primary outcome was reported measles incidence by age group during March 2018 through September 2019. Secondary outcomes were the national number of reported cases and the number and incidence reported for Jerusalem District [4,5]. The Netherlands, Czechia, and Australia were examined as structured comparisons using a predefined extraction framework rather than as equivalent quantitative validations. Across all settings, “subsequent measles activity” referred to the reported case burden or incidence observed after the immunity assessment, “importation” referred to infection acquired abroad or a transmission chain epidemiologically linked to an imported case, and “clustering” referred to a local or socially connected concentration of susceptible individuals or cases.

Published age-specific seropositivity was converted into a time-updated susceptibility profile. Subsequent activity was interpreted in relation to infectious introduction, susceptible density and distribution, contact structure, and control capacity. Susceptible density was defined conceptually as the number of susceptible individuals per geographic area or within a shared contact setting; it could not be estimated directly from national public data.

### Israel susceptibility reconstruction

We extracted the age-specific sample sizes and seropositivity estimates reported for a nationwide Israeli survey conducted in 2015 [3]. These published results were inputs to the present model; the original participant-level data were not used. Each 2015 age group was mapped to monthly birth cohorts and aged to 1 March 2018. Children born after the 2015 baseline were added as new monthly cohorts.

For cohort c in month t, the conceptual update was:

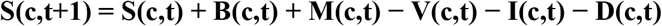

where S is the number susceptible, B represents births entering the cohort structure, M net migration, V effective immunization, I prior measles infection, and D deaths or aging out of the analytic group. In the public-data implementation, births and routine vaccination were represented explicitly; net migration, prior infection, and deaths were retained conceptually but could not be estimated by age and month and were therefore treated as unmeasured sources of uncertainty rather than assigned artificial values.

The population base used a Central Bureau of Statistics (CBS)-derived end-2015 national control total and a documented United Nations (UN) age-structure fallback. The first dose of measles-containing vaccine (MCV1) was applied from 12 months of age and the second dose (MCV2) from 6 years of age. For infants born after baseline, maternal antibody protection was allowed to decline before routine vaccination. Protection following vaccination was calculated as vaccination coverage multiplied by assumed vaccine effectiveness. For older cohorts already represented in the 2015 published results, agespecific seropositivity was carried forward without imposing additional unobserved waning. Three scenarios were prespecified to propagate uncertainty rather than to estimate statistical confidence intervals. The optimistic scenario combined the upper 95% confidence bound of the published seropositivity estimate with higher assumptions for protection among equivocal results, vaccination coverage, and vaccine effectiveness. The central scenario used the published point estimate, assumed that 50% of the aggregate equivocal fraction represented protection, and applied central vaccination coverage and effectiveness assumptions. The conservative scenario combined the lower 95% confidence bound with equivocal results treated as unprotected and lower vaccination assumptions. The resulting optimistic-to-conservative range is therefore a scenario-based uncertainty interval, not a 95% confidence interval for the modelled susceptible proportion. For age group a, the reported quantities were:

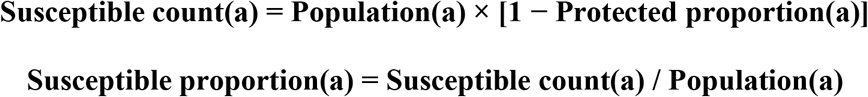

Because official single-age denominators, vaccination dates, migration by age, and subnational seropositivity were unavailable, the resulting totals were interpreted as scenario bounds, not as direct population-prevalence estimates.

### External comparisons

For each external setting, we extracted the published survey year, target population and sampling frame, age distribution, sample size, assay and positivity threshold, age-specific immunity results, definition and distribution of subsequent cases, vaccination status, imported or import-related status, and evidence of geographic or social concentration. The Netherlands was selected to examine whether high national seroprevalence could conceal dense susceptibility within a connected minority [6,7]. Czechia was selected to examine how sampling frame and age distribution influenced inferred immunity gaps [8,9,12]. Australia was selected to examine whether low detectable antibody predicted sustained transmission where most reported cases were imported or import related [10,11]. Estimates were compared descriptively and were not pooled because assays, thresholds, sampling frames, age groupings, and follow-up periods were not harmonized.

## Results

### Israel

At 1 March 2018, the modelled national population was approximately 8.59 million. The estimated number susceptible was approximately 545,000 (6.3%) under the optimistic scenario, 1.22 million (14.2%) under the central scenario, and 1.92 million (22.4%) under the conservative scenario. These values are model-derived national counts obtained by summing age-specific susceptible counts. The range primarily reflects alternative assumptions about equivocal serology, age-specific denominators, vaccination coverage, and vaccine effectiveness; it is a sensitivity range rather than three direct prevalence estimates.

Children aged <1 year had the highest central susceptible proportion, approximately 75.7% (scenario range, 68.4%–84.1%). This pattern was expected because maternally derived antibody had largely waned while routine MCV1 was not yet due. Children aged 1–4 years had the next-highest central susceptible proportion, approximately 27.3% (scenario range, 19.8%–36.6%), reflecting the transition from the pre-vaccination interval to incomplete or delayed first-dose protection. These were also the age groups with the highest published incidence: 196.0 per 100,000 among children aged <1 year and 118.9 per 100,000 among children aged 1–4 years (Figure 2) [3].

**Figure 1.**
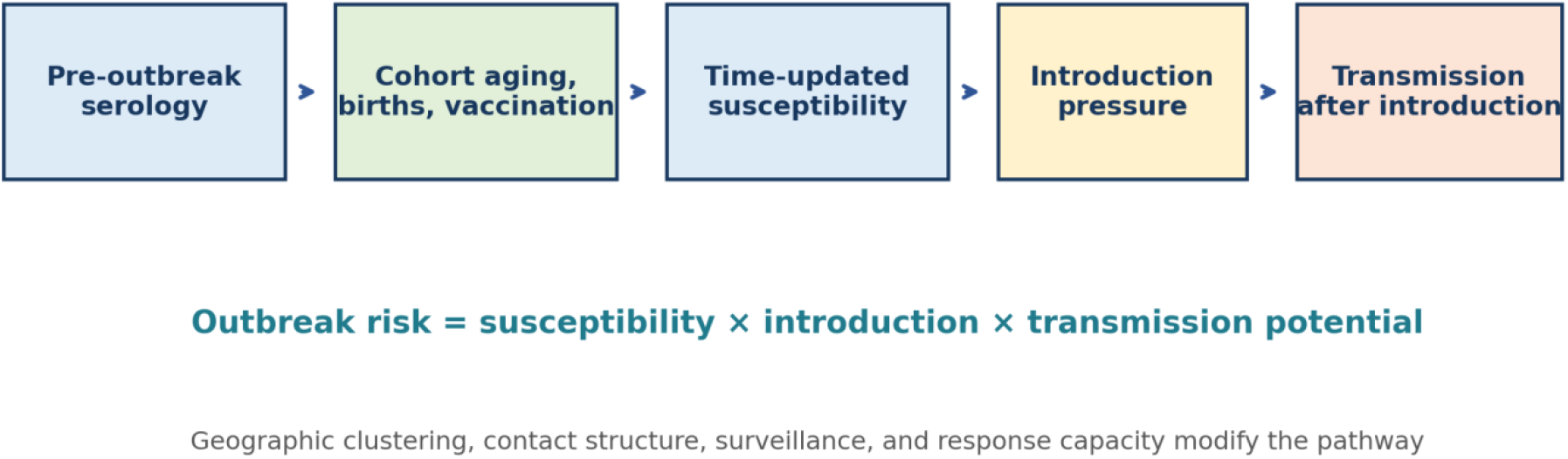
Analytical framework linking published seropositivity results to subsequent measles activity.

**Figure 2.**
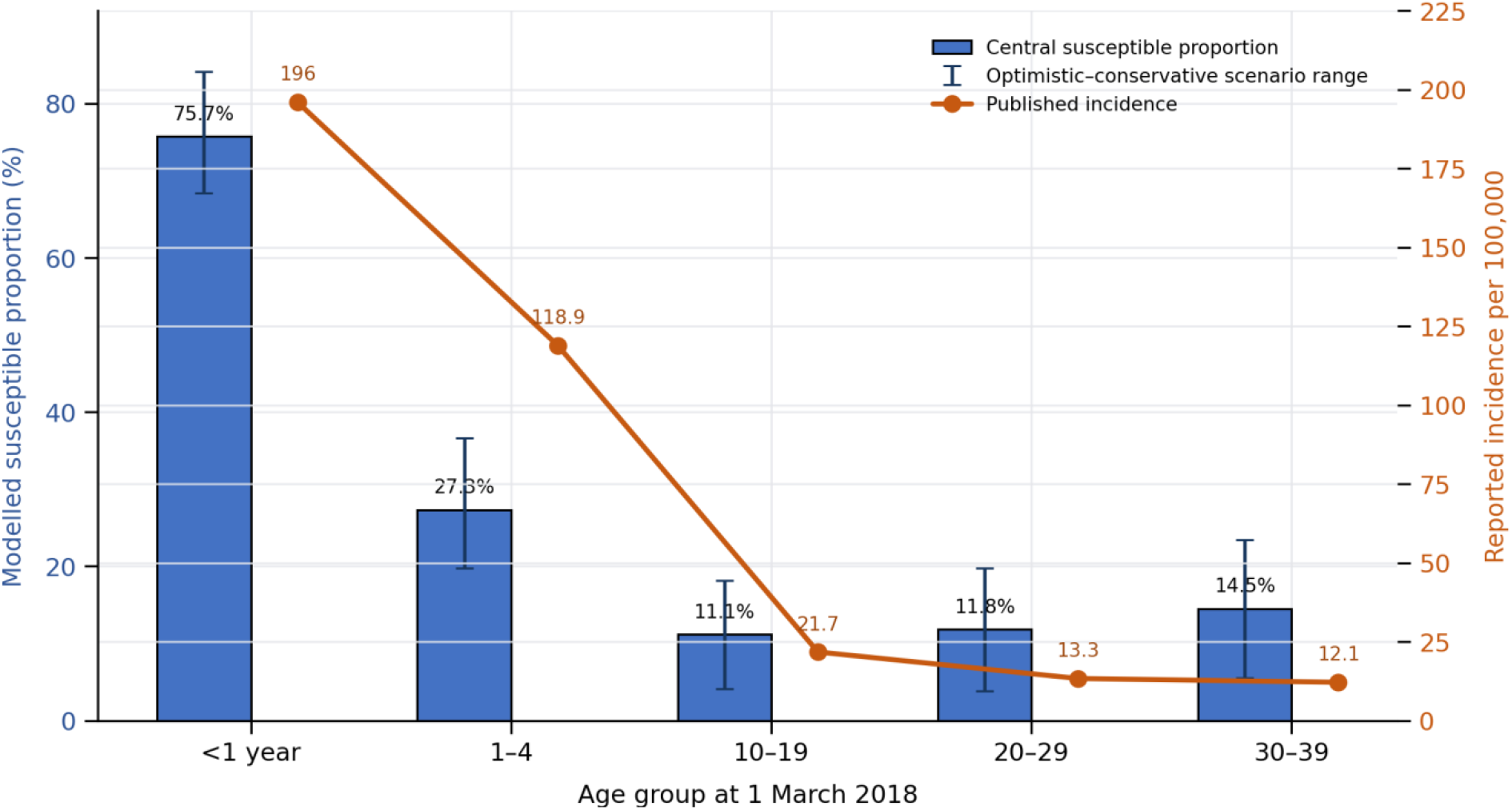
Modeled susceptible proportion on 1 March 2018 and subsequently reported measles incidence by age group, Israel.

Bars show the central modelled susceptible proportion; error bars show the optimistic-to-conservative scenario range and are not statistical 95% confidence intervals. Points show published national incidence during the 2018–2019 outbreak for the five age groups with extractable estimates [3]. Age-specific case numerators and denominators were not available in the published summary used for extraction; therefore, incidence confidence intervals were not calculated. The comparison is descriptive, and the model was not fitted to observed incidence.

The national age reconstruction did not, by itself, explain where transmission concentrated. Of 4,311 reported national cases, 2,202 occurred in Jerusalem District. First-dose coverage in heavily affected Orthodox communities was reported as 78.4%, compared with 90.1% in other communities [4,5]. This pattern supports the model rationale: a national susceptible count becomes more informative when the geographic and social concentration of susceptible people is also represented.

### Cross-setting comparison

Table 1 compares the four settings and shows that the same national seropositivity value can imply different epidemiological risks depending on clustering, exposure, and surveillance. In the Netherlands, 96% national seroprevalence concealed 51% susceptibility among Orthodox Protestant children younger than 10 years before a 2,700-case outbreak concentrated in that population [6,7]. In Czechia, age-specific gaps differed between a national survey and a separate adult study, illustrating sensitivity to survey design and sampled population [8,9]. In Australia, 80.8% EIA positivity and 8.9% equivocal results coexisted with sustained elimination, while 98.1% of notifications during 2012–2019 were imported or import related [10,11].

**Table 1.** Published pre-activity immunity signals, subsequent measles activity, and the model component highlighted in each setting.

| Setting | Published immunity signal | Subsequent activity | Interpretation for outbreak-risk modelling |
| --- | --- | --- | --- |
| Israel | 2015 overall seropositivity 90.7%; pronounced infant and young-child susceptibility [3] | 4,311 national cases; 2,202 in Jerusalem District [4,5] | Age-specific susceptibility was informative, but local coverage and clustering were needed to explain concentration. |
| Netherlands | 2006–2007 national seroprevalence 96%; 51% susceptible among Orthodox Protestant children <10 years [6] | 2,700 reported cases; 94% unvaccinated; concentration in Orthodox Protestant communities [7] | Subpopulation denominators, local coverage, and network connectivity were more informative than the national average. |
| Czechia | 2013 national seroprevalence 93%; 62% at age 1 and 80.4% at ages 35–44; lower values in a separate adult study [8,9] | Outbreak activity during 2017–2019; most cases in a later 2018–2024 summary occurred in 2019 [12] | Survey design, assay interpretation, age-specific outcomes, importation, and genomics need reconciliation. |
| Australia | 2012–2013 EIA positivity 80.8% and equivocal results 8.9%; declining detectable antibody in ages 10–39 [10] | 1,337 notifications during 2012–2019; 98.1% imported or import related [11] | Introduction pressure, vaccination status, response capacity, and limited onward transmission changed the meaning of low detectable antibody. |

## Discussion

The findings indicate that seropositivity is an informative measure of population susceptibility but is insufficient, on its own, to characterize outbreak risk. Sustained transmission requires both an infectious introduction and a sufficiently large, dense, and interconnected susceptible population. For a pathogen with the transmission potential of measles, modest national immunity gaps may have disproportionate consequences when susceptible individuals are concentrated within the same locality or contact network.

The Israeli reconstruction provides a transparent example. Aging published 2015 results to the 2018 threshold recovered the expected vulnerability of infants and young children, and those groups subsequently had the highest reported incidence. The result is epidemiologically plausible rather than surprising: infants pass through a predictable interval after maternal antibodies wane and before routine vaccination. The Jerusalem experience then shows why an age-only national model is insufficient. Susceptibility was not randomly distributed; it was concentrated in communities with lower coverage and close contact patterns [4,5].

These findings also have direct relevance to Israel’s updated immunization schedule. Although the present analysis did not simulate alternative vaccination schedules, the substantial susceptibility estimated among young children supports the epidemiological rationale for advancing the second routine MMRV dose from 6 years to 18 months of age. The revised schedule, implemented in Israel from 1 July 2026, shortens the interval during which children rely on protection from a single dose and provides an earlier opportunity to complete the two-dose series. This consideration is particularly relevant in densely populated communities and in large households, where repeated close contact can amplify transmission when vaccination is delayed or incomplete. The policy change is particularly timely in the context of the prolonged 2025–2026 outbreak, which again demonstrated how delayed or incomplete vaccination can sustain transmission in densely populated communities with large numbers of young children.

The external settings sharpen the same point. The Dutch national average hid a connected minority with very high susceptibility [6,7]. Czech estimates changed with the sampled population and survey design [8,9]. In Australia, low detectable antibody did not translate into endemic transmission because most activity followed importations and public-health response limited onward spread [10,11]. Together, these examples support a model with three explicit layers: susceptible population, introduction pressure, and transmission conditions.

This study remains a proof of concept. The Israeli totals depend on public demographic allocation and vaccination assumptions, and the external case studies were not reconstructed with the same level of detail. The model does not yet estimate individual risk or prospective outbreak probability. Its value at this stage is to define the minimum information needed for a later validated model and to show why a single national coverage or seropositivity value is inadequate.

## Conclusion

Published age-specific seropositivity estimates can be translated into a time-updated susceptibility profile, but the profile becomes epidemiologically useful only when it is linked to where susceptible people are concentrated, whether measles is introduced, and whether local contact and response conditions permit sustained transmission. The next validation step is a harmonized analysis using official single-age denominators, vaccination timing, sub-national coverage, importation data, and time-aligned outcomes in several settings.

## Data availability statement

All analyses use published aggregate results and public demographic information. The formula-based Israel model and standardized multicountry extraction workbook are intended as supplementary methodological files (available upon request). No original participant-level serosurvey data or restricted individual-level surveillance data were accessed.

## Ethics statement

This public-data proof of concept used published aggregate information only. Institutional confirmation that formal ethics review was not required should be obtained before submission.

## Author contributions

Eran Kopel: Conceptualization, methodology, formal analysis, writing original draft, and visualization. Ravit Bassal: Methodology, interpretation, validation, review and editing.

## Acknowledgements

None.

## Use of artificial intelligence tools

Microsoft 365 Copilot, based on the GPT-5 reasoning model accessed through Microsoft Edge, was used during July 2026 to assist with English-language drafting and editing, manuscript organization, and the development of code used to implement the authors’ prespecified formula-based calculations and generate the study figures. The tool did not generate or supply original study data. All model assumptions, calculations, numerical results, figures, references, interpretations, and policy conclusions were reviewed and verified by the authors, who take full responsibility for the accuracy and integrity of the final manuscript.

